# A Method for Machine Learning Generation of Realistic Synthetic Datasets for Validating Healthcare Applications

**DOI:** 10.1101/2021.02.11.21250741

**Authors:** Theodoros N. Arvanitis, Sean White, Stuart Harrison, Rupert Chaplin, George Despotou

## Abstract

**Background:** Digital health applications can improve quality and effectiveness of healthcare, by offering a number of tools to patients, professionals, and the healthcare system. Introduction of new technologies is not without risk, and digital health applications are often considered a medical device. Assuring their safe operation requires, amongst others, clinical validation, which needs large datasets to test their application in realistic clinical scenarios. Access to such datasets is challenging, due to concerns about patient privacy. Development of synthetic datasets, which will be sufficiently realistic to test digital applications, is seen as a potential alternative, enabling their deployment.

**Objective:** The aim of work was to develop a method for the generation of realistic synthetic datasets, statistically equivalent to real clinical datasets, and demonstrate that Generative Adversarial Network based approach is fit for purpose.

**Method:** A generative adversarial network was implemented and trained, in a series of six experiments, using numerical and categorical variables from three clinically relevant datasets, including ICD-9 and laboratory codes from the MIMIC III dataset. A number of contextual steps provided the success criteria for the synthetic dataset.

**Results:** The approach created a synthetic dataset that exhibits very similar statistical characteristics with the real dataset. Pairwise association of variables is very similar. A high degree of Jaccard similarity and a successful K-S test further support this.

**Conclusions:** The proof of concept of generating realistic synthetic datasets was successful, with the approach showing promise for further work.

## INTRODUCTION

Digital health has seen a continuously increasing number of innovative applications, aiming to improve all aspects of one’s health and care; such as safety, efficacy and monitoring of care plans, empowerment of the patient to manage their own condition, as well as discovery of new clinical knowledge. These innovations have been made possible by applying state-of-the-art computer science, data science, and software engineering technologies in healthcare, resulting in applications such as automated diagnosis, self-monitoring, telehealth and clinical decision support. Applications can vary from simple statistics viewers, to symptom checkers and diagnostic services, used in both primary and secondary care. Their integration with clinical pathways can be seen as introducing Clinical Decision Support (CDS). Failures in their operation may cause harm to patients. For example, by offering incorrect advice, by making the wrong diagnosis, or recommendation to a healthcare professional. This potential for harm is increasingly recognised and has been incorporated in regulation, where software is seen as a medical device [1,2,3]. Software applications may expose patients to risks, due to unintended or erroneous behaviour, or lack of clinical validation that may create unknowns, in whether these applications are fit-for-purpose and acceptably effective. Understanding risks and establishing clinical validation is necessary, for software manufacturers to be able to meet certification and regulation requirements, in order to offer their products to the healthcare system and patients. Clinical validation entails comparing the application against datasets, typical of the patients who will use it. This can be problematic as large patient datasets are not readily available to manufacturers due to regulations and law governing patient privacy [4]. The problem is aggravated, as datasets need to be suitable for the context of an application, with manufacturers often resorting to buying compiled data. An increasing number of applications requiring datasets also increases demand and hence waiting times. Furthermore, even if manufacturers can access data, either through a source or produced by themselves, validation is still difficult as the regulator will need to have access to the same dataset (or a common dataset based on which to exchange information about the fitness of the application). As a result, patients may be deprived of digitally enabled applications improving the healthcare quality they receive. Use of Realistic Synthetic Datasets (RSDs) is seen as a promising solution [5, 6, 7, 8, 11], addressing privacy challenges, whilst overcoming issues with alternatives; such as anonymized data that may skew the results of validation due to missing fields [4]. Natural Language Processing can enhance datasets by adding synthetic clinical notes, based on real records [9, 10]. RSDs consist of software-generated data points, which overall demonstrate equivalent statistical properties as a real clinical dataset. In the context of clinical validation, use of the two datasets should result in the same conclusions, with the same confidence. Contrary to using de-identified or anonymized datasets, RSDs: 1) do not need prolonged preparation and approval process, something that is required even with anonymized data; 2) can include variables that may be considered sensitive with respect to patients’ privacy and are not included in anonymized and de-identified datasets; 3) are highly resistant to cross-referencing with other datasets (although some concerns still remain to be addressed). The most common approach, to developing synthetic datasets, looks at the associations and the distributions of variables of interest, and develops probabilistic models that will then generate the synthetic data [12-15]. A potential drawback of this is that they often use a driving variable, which is generated first, and then drives the associations with the others [8], potentially skewing the dataset to conditions represented by these variables, and omitting other variables that may hide unearthed associations.

Machine Learning (ML) has been prominent in producing large RSDs, with similar statistical qualities to a real dataset on which they are trained [16, 17, 18]. Generative Adversarial Networks (GANs) are a ML approach based on neural networks, recently recognized as able to accurately mimic real datasets [19, 20, 21, 22]. GANs, in general, will start their training without relying on statistical models representing real dataset, and will establish the statistical properties of the real dataset [23]. This allows GANs to be ‘dataset agnostic’ and transferable across multiple datasets. Evaluating the performance of GANs requires focusing on statistical properties of the real and the synthetic dataset [24, 25]. This paper presents the results of a Realistic Synthetic Dataset Generation Method (RSDGM) using GANs, which generated a synthetic dataset suitable for validation of digital health applications.

## OBJECTIVES

A proof of concept of producing realistic, large-scale, scalable, machine-learning generated datasets, for validation of healthcare applications used across all levels of care, including:

- *Realistic*: the dataset will need to be statistically equivalent to real dataset.
- *Large-scale*: the dataset will need to consist of a large number of entries and variables.
*Scalable*: the method should be scalable for larger datasets.
- *Machine learning generated*: machine-learning (GANs) should be used to generate the dataset, and the optimum hyper-parameters need to be identified.
- *Validation of healthcare applications*: the method should generate evidence justifying the suitability of the RSD to validate healthcare applications.
- *Levels of care:* RSDs for primary as well as secondary care.

## METHOD

The method (Figure 1) consists of four (4) main steps (1, 2, 3, and 4) and three (3) contextual steps (a, b, and c). The contextual steps provided the necessary framework for making methodological decisions. The method was defined iteratively over six main experiments (Table 1), each of which consisted of numerous runs, which allowed testing aspects such as the GAN hyper-parameters. For example, changing the hidden layers twice in experiment #3, which tested three different GANs in parallel, would result in six runs. All experiments were run on a computer with 2x Intel Xeon Gold 6144 3.5GHz, 3x Nvidia Quadro RTX5000, 8×16GB DDR4 2666MHz RDIMM ECC, and Windows 10 Professional for workstations.

**Table 1.** Overview of the six experiments; the dataset used, the type of GAN implemented, the complexity added in each experiment and the approximate duration of each run in the experiment.

| Exp # | Dataset | Type of GAN | Additional Complexity | Appx. Duration of a Run |
| --- | --- | --- | --- | --- |
| 1 | HES | GAN | Simple GAN, Numerical values. | 10 mins |
| 2 | MIMIC-III | GAN, CGAN, WGAN | Multiple GAN implementations. Increase in epochs. | 25 mins |
| 3 | MIMIC-III | GAN, CGAN, WGANGP | Multiple GAN implementations, Increase in dataset, categorical values. | 45 mins |
| 4 | A&E | GAN, CGAN, WGANGP | Dataset transferability | 20 mins |
| 5 | MIMIC-III | WGANGP | ICD-9 codes. | 8 hrs |
| 6 | MIMIC-III | WGANGP | Lab test codes, and association with ICD-9 codes. | 12 hrs |

**Figure 1.**
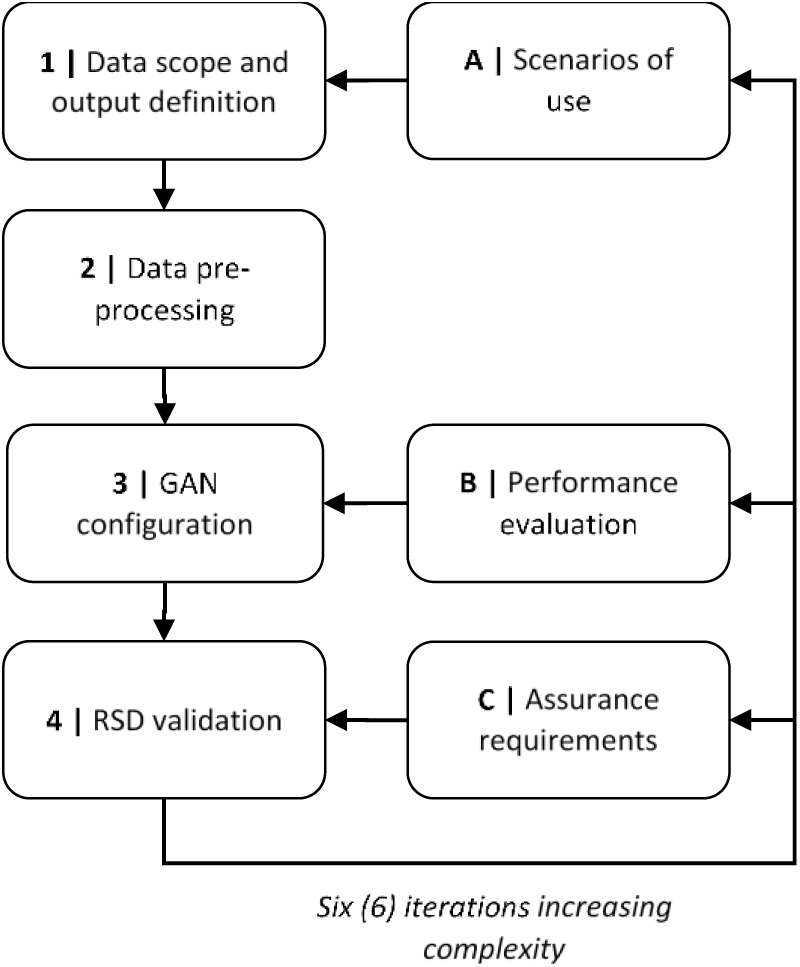
Overview of the Realistic Synthetic Dataset Generation Method (RSDGM)

### 1. Data Scope and Output Definition

This step defined the content and purpose of the dataset, providing the business goals for the experiments, and specifies the success requirements of the RSD:

- The method needs to be easily scalable to re-train the dataset with additional variables.
- Allow modelling of large number of variables.
- Establish associations amongst all variables.
- Produce both numerical and categorical values.
- Allow the generation of clinical codes (e.g., ICD-9).

Table 1 shows an overview of the experiments performed as part of developing the RSDGM. Three different datasets (NHS HES [26], NHS A&E [27]. and MIMIC III [28]) were used to ensure the transferability of the method. MIMIC III was considered the most complete dataset, and being open, was preferred for the final experiments.

Experiment #1 focused on a vanilla GAN, with numerical values. Experiments #2, #3, #4 compared the implementations of three different GANs; a vanilla GAN (GAN) [30], a conditional GAN (CGAN) [31], and a Wasserstein GAN (WGAN) [32], subsequently implementing gradient penalty (WGANGP) [33]. The WGANGP was considered to have the best performance, and was selected for the last experiments, but with the added complexity of generating ICD-9 codes (exp. #5), and lab test codes (exp. #6).

#### Context A: Scenarios of use

The main scenario was the validation of clinical decision support applications, providing recommendations to patients or healthcare professionals [29]. The scope encompassed both primary and secondary care. Primary care datasets focus on a relatively low number of variables (e.g., weight, age, diagnosis) generally describing bigger population. Secondary care datasets need a larger number of variables, relating to the protocol followed during the encounter of the patient, including lab tests.

### 2. Data Pre-processing

Pre-processing removed missing and duplicate data. Multiple categorical features were interpreted as a collection of binary features, with one-hot encoding. The input to this transformer is a matrix of integers, denoting the values of the categorical features. The output is a sparse matrix, where each column corresponds to one value of a feature. All variables of the training data are rescaled by applying standardisation. We multiply the forecast value of the standardised input by the standard deviation calculated in the original series, and then add the mean. Label encoder was used for the categorical variables to generate their combined pairwise correlations.

There are two main challenges in incorporating lab codes and ICD-9 variables in the generation method. The first is the complexity of the variables. There are thousands of ICD9 codes in the dataset, providing a large amount of values for the GAN to synthesize. Furthermore, the diagnosis variable (where the ICD-9 codes are found) is a composite variable, consisting of multiple ICD-9 codes in simple text format. The second is that the method needs to capture the associations between the individual ICD-9 codes (e.g., co-morbidities of a patient), as well as the combination of ICD-9 codes with the patient demographics. In order to achieve an approach that is truly agnostic to the dataset, the method should not receive declared associations between these variables, but instead the GAN discern them during the learning process.

In the experiments, we generated ICD-9 codes per hospital admission, using the DIAGNOSIS_ICD.csv file of MIMIC-III. In this, each patient has multiple hospital stays and each stay is associated with a unique ICD-9 code. The diagnosis variable was expanded in a separate matrix with length equal to the number of unique codes, containing binary values capturing the presence or not of an ICD-9 code (Table 2).

**Table 2.**
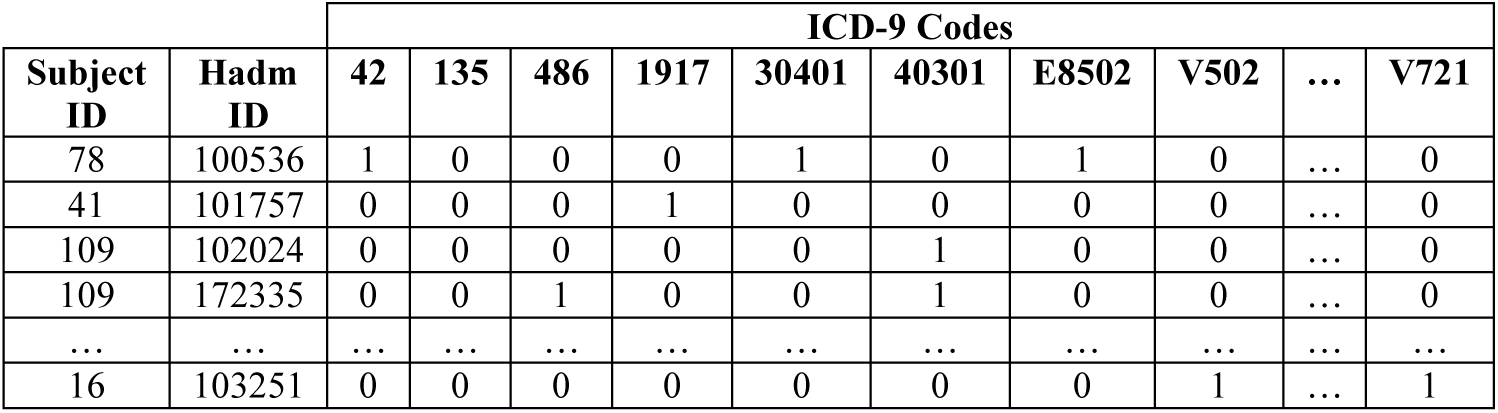
Excerpt of patient – ICD9 code matrix

| Subject ID | Hadm ID | ICD-9 Codes |  |  |  |  |  |  |  |  |  |
| --- | --- | --- | --- | --- | --- | --- | --- | --- | --- | --- | --- |
|  |  | 42 | 135 | 486 | 1917 | 30401 | 40301 | E8502 | V502 | ... | V721 |
| 78 | 100536 | 1 | 0 | 0 | 0 | 1 | 0 | 1 | 0 | ... | 0 |
| 41 | 101757 | 0 | 0 | 0 | 1 | 0 | 0 | 0 | 0 | ... | 0 |
| 109 | 102024 | 0 | 0 | 0 | 0 | 0 | 1 | 0 | 0 | ... | 0 |
| 109 | 172335 | 0 | 0 | 1 | 0 | 0 | 1 | 0 | 0 | ... | 0 |
| ... | ... | ... | ... | ... | ... | ... | ... | ... | ... | ... | ... |
| 16 | 103251 | 0 | 0 | 0 | 0 | 0 | 0 | 0 | 1 | ... | 1 |

Each patient has a unique id (‘SUBJECT_ID’) and each is associated with a unique hospital admission id (‘HADM_ID’).

### 3. GAN Design and Configuration

Table 3 presents the configurations for the GANs, in each experiment. In experiments #2, #3, and #4 the same configuration was used for all three GANs.

**Table 3.** GAN configurations for each experiment

| E# | Epochs | Learning rate | Hidden layers | Neurons | Batch size | Optimiser |
| --- | --- | --- | --- | --- | --- | --- |
| 1 | 10,000 | 1e-3 | 3 D, 2 G | D[8, 8, 8, 1]<br>G[8, 8] | 150 | RMSProbOptimiser |
| 2 | 30,000 | 1e-4 | 1 D, 1 G | D[18,1]<br>G[21] | 130 | Adam |
| 3 | 50,000 | 1e-4 | 1 D, 1G | D[18,1]<br>G[21] | 5000 | Adam |
| 4 | 10,000 | 1e-4 | 1 D, 1 G | D[18,1]<br>G[21] | 1000 | Adam |
| 5 | 30,000 | 1e-4 | 2 D, 2 G | D[256,128,1]<br>G[256, 128] | 150* | Adam |
| 6 | 50,000 | 1e-4 | 2 D, 2 G | D[256,128,1]<br>G[256, 128] | 379* | Adam |

The Adam optimiser [34] showed best performance in early evaluation. In the last experiments, two hidden layers, both for the discriminator (D) and the generator (G), were used. The number of neurons was the *sum of input and output layers multiplied by 2/3 of the value given between the input and output sizes*. Mean squared loss for numerical variables (e.g., age) and cross entropy loss for categorical variables were initially used. However, Wasserstein loss was later used, improving the results. The first experiment used the Leaky ReLu activation function, whereas subsequent experiments used Rectified Linear Units (RELU), as the activation function of the Generator, except of the output layer where *tanh* was used for numerical variables. For the discriminator, ReLU was used for all activation functions, except for the output layer, where the sigmoid function for categorical and *tanh* for numeric features were used [35]. The number of epochs ranged from 10,000 to 50,000. A trial implementation tested the network up to 300,000 epochs, showing that, after 50,000 epochs, changes were insignificant. A target of 50,000 epochs was considered a good balance between training and computing power. The number of discriminator iterations per generator iteration is 10, and the Gradient Penalty (GP) (*lambda-penalty coefficient*) is 10. The raw generated data values were continuous in range 0 to 1, converted to binary (0 or 1) through rounding for categorical data classification. For numeric features, the output of generator is de-standardised.

#### Context B: Performance evaluation

As this was an exploratory, proof-of-concept study, the method evolved along with the results of the experiments. The performance evaluation, used for this, was a meta-process. It analysed the results and shaped the final form of the experiments in Table 3. After each experiment, this approach identified parameters for rapid trial and error, as well as the improvements in subsequent experiments (e.g., increase of epochs).

### 4. Synthetic Dataset Validation

This step examined whether the RSD would be fit for purpose for the objective of the study and the scenario identified in context A. From the early stages of the experiment, it was clear that the validation of the synthetic dataset is a major challenge. Although the dataset contains different data points, its overall qualities needed to be equivalent to the real dataset. Qualifying the justification for equivalence requires understanding the validation context, in order to identify suitable evidence. This was achieved by a parallel, assurance process (Context C). A justification outline was developed, identifying the evidence needed to be generated in order to support it [36]. The aspects needed support by evidence were recognised by conducting a safety assessment, identifying potential risks using the RSDGM. The identified justification comprises of three main arguments. Firstly evidence needs to demonstrate that the RSD is a high fidelity representation of clinical knowledge (e.g., prevalence of conditions), including potential relationships not represented in existing knowledge. Secondly, there needs to be evidence supporting that the synthetic and the real datasets exhibit almost identical statistical properties (e.g., associations amongst variables). The final argument of the justification focuses on the technical correctness and appropriate application of the generation method. The first argument is beyond the scope of this paper, as it focuses on the use of the RSD. The second and third arguments, resulted in a process identifying evidence, which would convincingly justify the validation of the RSD. The loss function, as well as scatter plots of the datasets through regular intervals of training the GAN, demonstrate that the network performed as predicted by theory. Visual comparison of the data entries, association tables, jaccard similarity, and a K-S test, were identified as convincing evidence to support statistical equivalence.

## RESULTS

### Comparison of the GAN Implementations

Wasserstein loss consistently produced the best results in the experiments, where multiple GANs were implemented. Figure 2 presents the results (selected variables) of experiment #3, where 10 features were selected from the ADMISSION, PATIENT and ICUSTAYS tables of the MIMIC-III dataset; 7 were categorical and 3 numeric, trained for 50,000 epochs. The scatter plots show the real data (points in blue) overlapped with the generated data (points in orange).

**Figure 2 (bottom).**
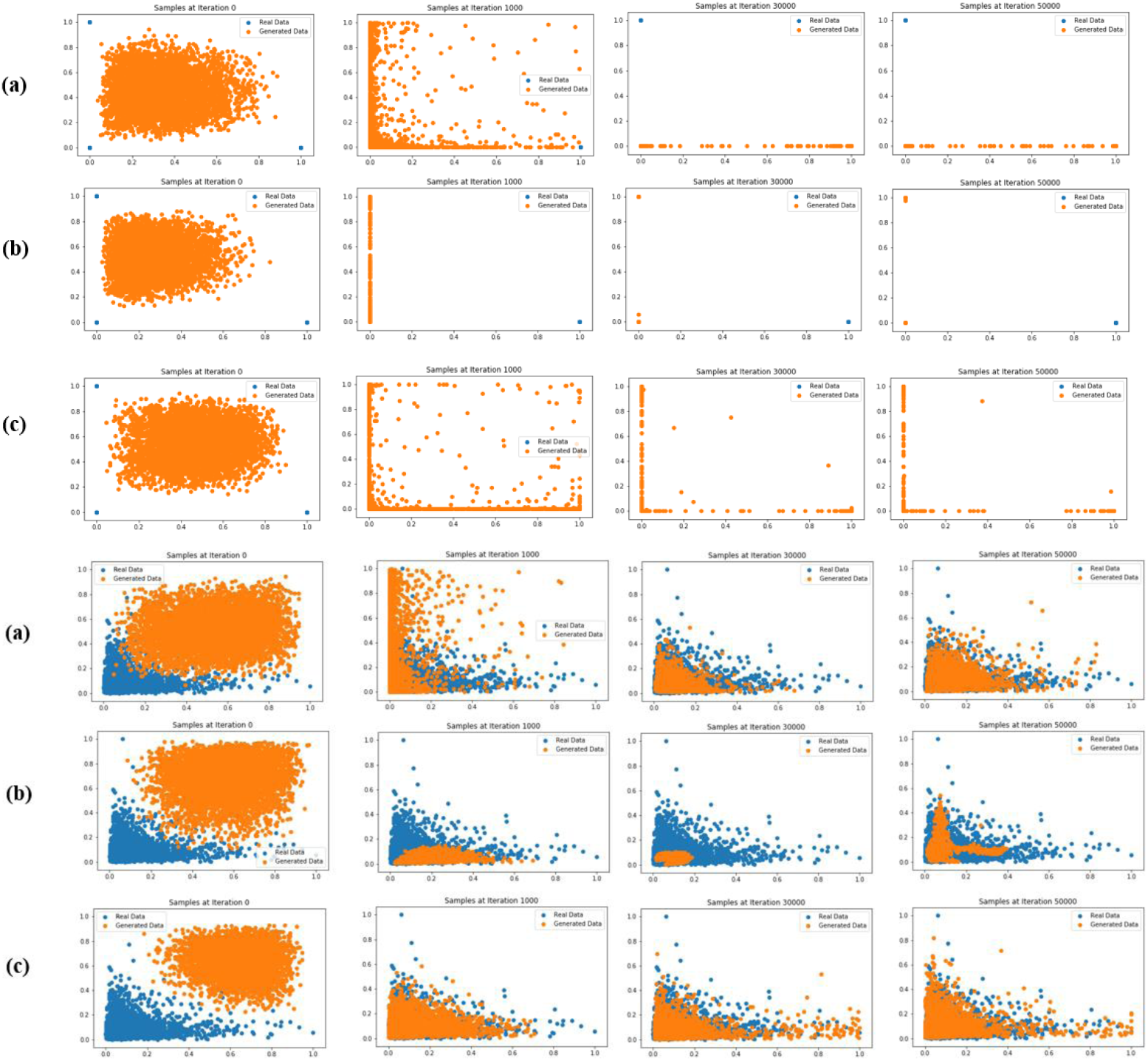
Scatter plots of synthetic data generation process for two selected numerical features for 50,000 epochs: (a) GAN, (b) CGAN and (c) WGANGP; (top) – Scatter plots of synthetic data generation process for two selected categorical features for 50,000 epochs: (a) GAN, (b) CGAN and (c) WGANGP

The WGAN performed best with the two types of variables. The vanilla GAN did not manage to perform well generating categorical values completely missing one dimension, whereas the CGAN was very poor generating numerical values.

Figure 3 presents the losses functions (10 iteration intervals; Generator in blue and Discriminator in orange). The WGAN implementation gave a better loss function plot with the Generator achieving good response, and the Discriminator not able to confidently discern true and false data.

**Figure 3.**
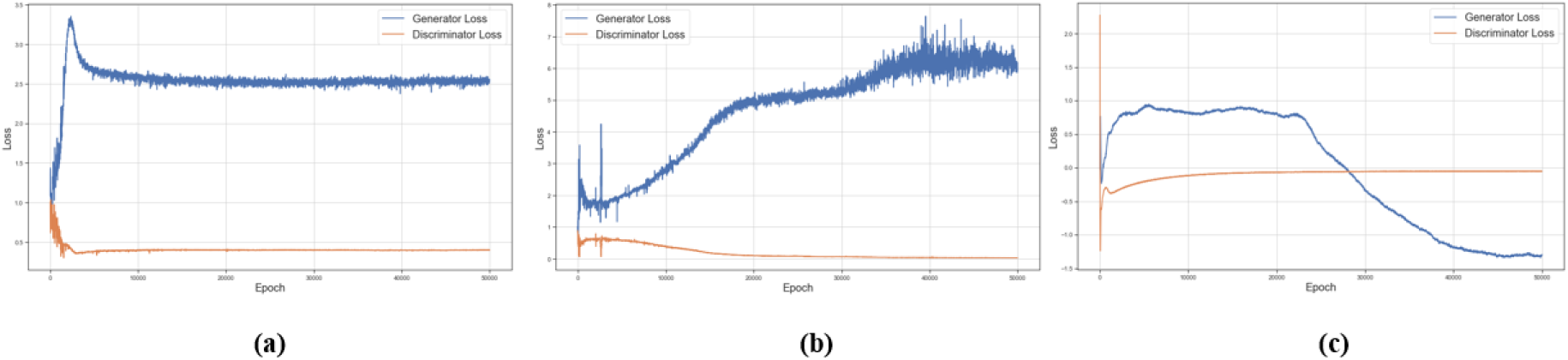
Generator and Discriminator losses for 50,000 epochs: (a) GAN, (b) CGAN and (c)WGAN

### RSDGM Validation framework

The validation approach of the RSDGM consists of a) loss analysis, b) visualisation of real and generated data, c) correlations comparison, and d) similarity measurement. The loss functions of the discriminator and generator were checked for two common problems: a) the generator collapsing and generating only one sample, and b) the generator simply memorizing or being too similar to the training data. The discriminator loss is trying to minimize the classification error of the discriminator and generator loss is trying to maximize the classification error of the discriminator. Although the loss function is not providing direct evidence about the statistical equivalence of the two datasets, it was particularly useful in early experiments, as it offered confidence that the network was operating as intended. It offered some early evidence and confidence about the algorithmic determinism of the implementation.

Visualisation of data was the first type of evidence produced, able to demonstrate statistical equivalence. It offers an intuitive way to understand the datasets, particularly effective in smaller size experiments, as it gave sufficient confidence about the similarity of the datasets; whilst giving the opportunity to spot potential outliers or over/under fitting. Furthermore, the plots gave an understanding of the behaviour of the network during its training, being plotted every 1000 iterations. They visualise how the Generator network starts, with a random initial mapping between the input and dataset vector space, and then gradually evolves to resemble the real dataset.

A correlation matrix of each dataset was computed in each experiment. The correlation matrix was visualized as a heat map. Correlation matrices provide confidence that the synthetic dataset has maintained an equivalent association amongst variables. Pearson correlation was applied for count or label-encoded categorical features. Spearman correlation was applied for binary features, as well as for experiments with mixed variables.

Ensuring whether the RSD learned the distribution of each dimension acceptably, the Mann-Whitney U test was applied. The test was used to compare whether the distributions of each variable of real and generated dataset come from the same population. Furthermore, two-sample Kolmogorov-Simirnov (K-S) test, which is a non-parametric test that compares the cumulative distributions of two data sets, was used to compare real and generated datasets. The null hypothesis of this test is that both samples originate from a population with the same distribution.

Finally, Jaccard similarity indices were used to compare associations, limited to absence/presence data. It is a measure of similarity for the two sets of data, with a range from 0% to 100%. The higher the percentage, the more similar the two populations. We considered dichotomic variables with 0 or 1 values, absence – presence of ICD9-codes or Lab item code per patient admission, and calculate the similarities between real and generated data for each code.

### WPGAN Generated Realistic Synthetic Dataset

The RSDGM used the MIMIC III dataset [37] as the real dataset to train on. The experiment (#6) that resulted in the final generated dataset was run using the following hyper-parameters: a) 50,000 epochs, b) a ReLU activation function, c) learning rate of 10^−4^, d) 2 hidden layers D [256,128,1] G [256, 128], Wasserstein distance as loss function, f) the Adam optimized, and g) penalty gradient 10. The experiment used the DIAGNOSIS_ICD and LABEVENTS tables of the MIMIC-III dataset. In total, 1357 codes (944 ICD9 + 413 Lab item) and 379 common unique hospital admissions of 300 patients were used as the real dataset.

Figure 4 illustrates the distribution of a sample of the variables in the RSD and the real dataset. The synthetic dataset shows a good representation of both numerical and categorical datasets with very similar distributions. The loss function (not illustrated) followed the expected response, similar to Figure 3 (right).

**Figure 4.**
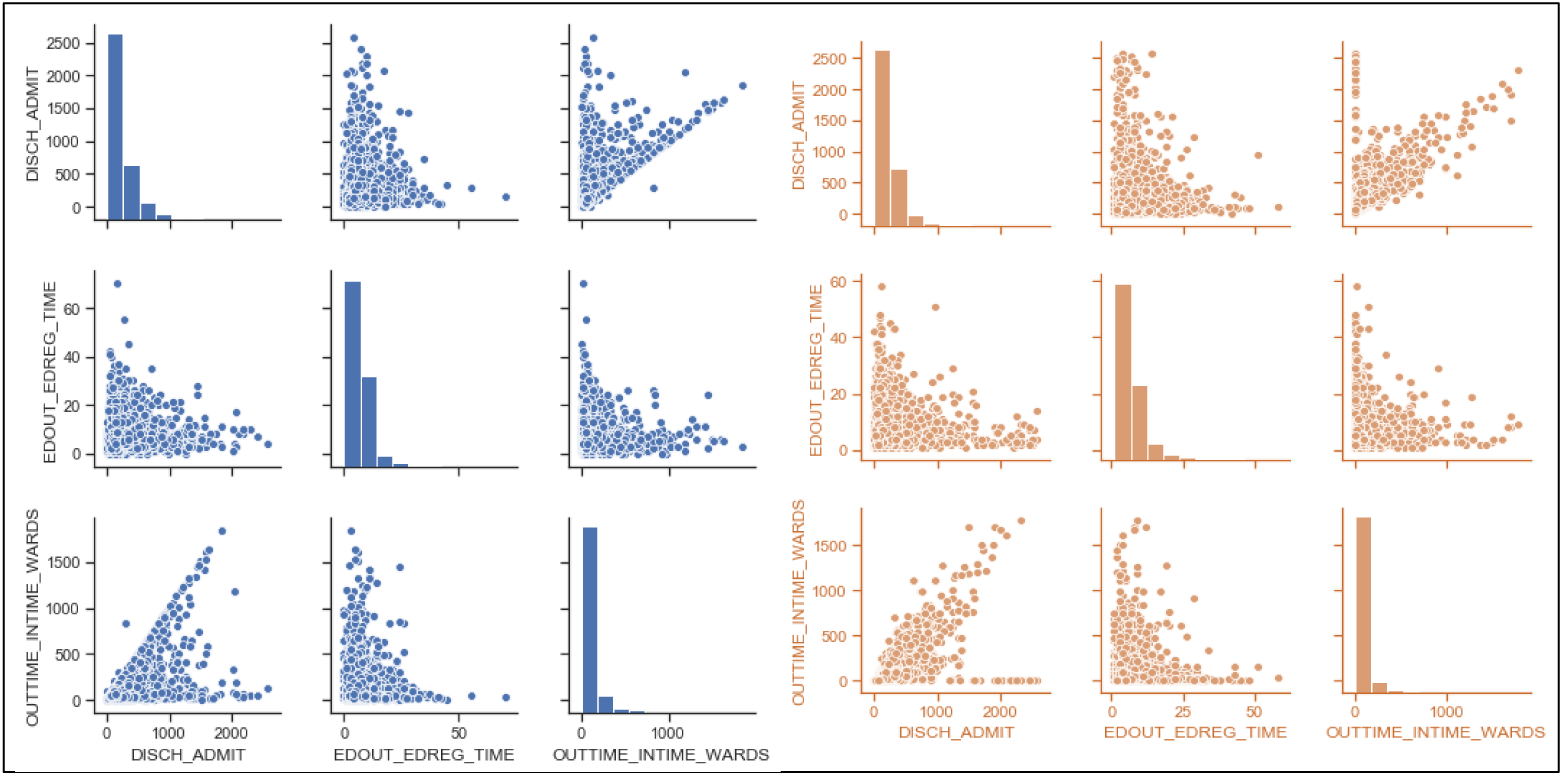
Distribution of a selection of numerical values in the two datasets (blue: real, orange: synthetic)

Figure 5 shows an extract of matrices of pairwise Spearman correlation of variables in the RSD and the real dataset. The matrices contain an extract of ICD-9 and lab codes. The very large dimensions of the full matrix do not allow complete graphical representation; nevertheless, further sample association matrices replicated the behaviour, also confirmed by a manual inspection of the associations. The RSD has preserved the associations between ICD-9 codes and the lab codes, of the original dataset.

**Figure 5.**
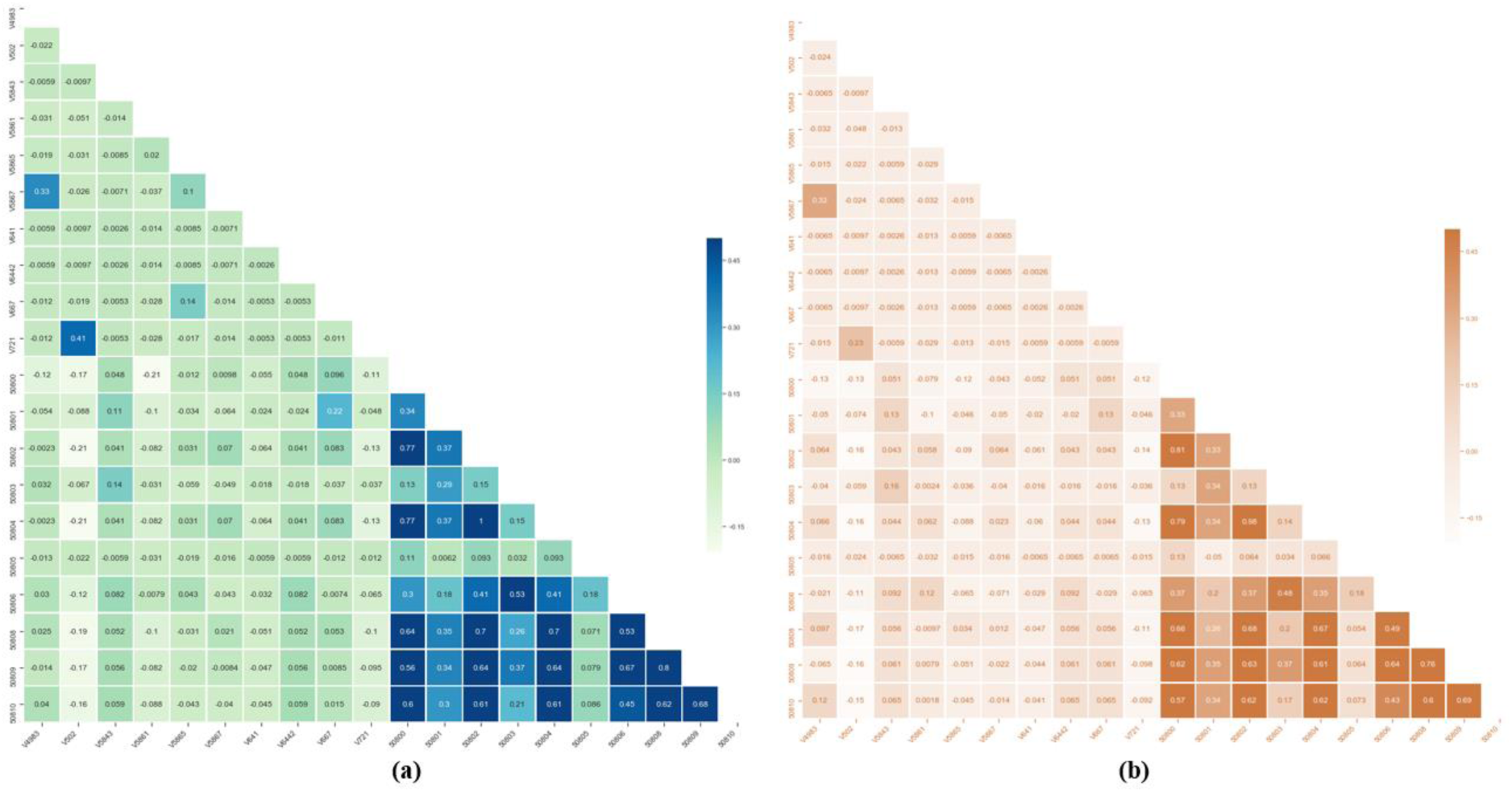
Correlation matrices of (a) real and (b) generated data for 10 ICD9-codes and 10 Lab item code

Figure 6 (left) shows the Jaccard similarity of the ICD-9 and lab item codes. The majority of the variables indicated very high Jaccard similarity, and only a fewer lab codes resulted in low similarity. A K-S test was performed with p = 0.05 examining whether the real and synthetic data samples were a subset of the same population, failing to reject the null hypothesis.

**Figure 6 (left).**
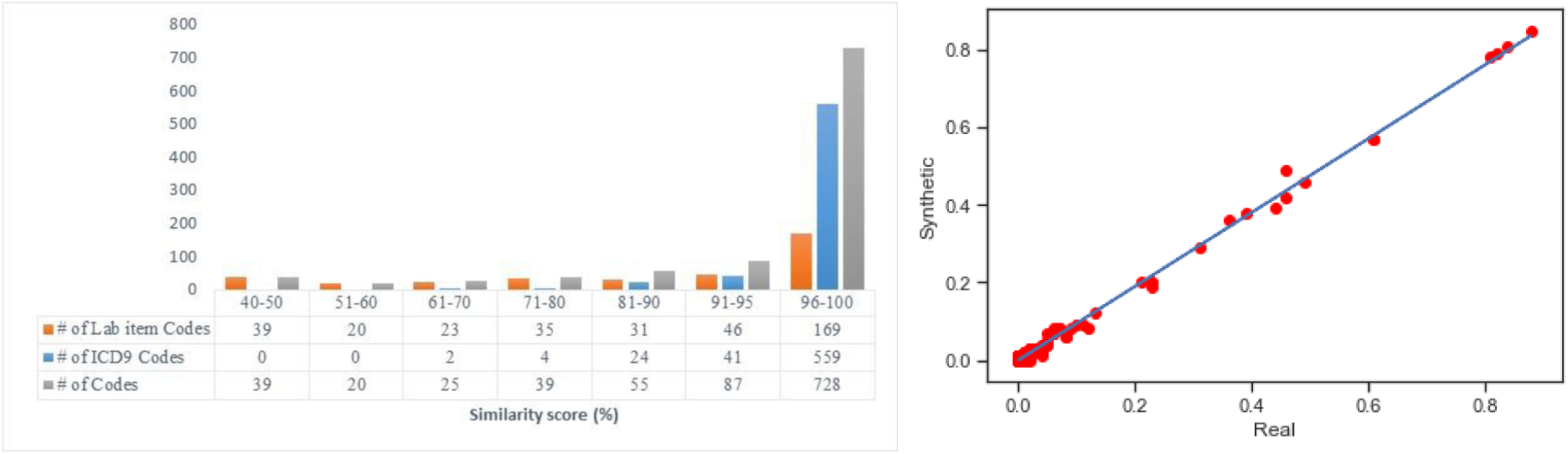
Histogram of Jaccard scores for lab item codes, icd9 codes and total number of codes; Figure 6 (right) Scatter plot of dimension-wise probability results of real binary data (x-axis) vs. synthetic counterpart (y-axis) produced

The scatter plot of dimension-wise probability results of real binary data vs synthetic counterpart is shown in Figure 6 (right). The K-S test failed to reject the null hypothesis with p-value (α = 0.05). Thus, there is no significant difference between the distributions for the two samples. Experiment six, being the final experiment, provided evidence towards the proof of concept of the RSDGM, using a highly complex dataset, with complex associations.

## DISCUSSION AND CONCLUSIONS

Realistic synthetic datasets are an approach recognised as promising, for validation and safety assurance of intelligent healthcare applications. This will overcome barriers of using datasets due to privacy concerns, enabling development of applications that may increase patient benefit. The GAN based method, successfully generated a realistic synthetic dataset. Statistical tests demonstrated that the two datasets share very similar qualities. Some differences between the datasets were identified, particularly with respect to certain lab and ICD-9 codes. This was attributed to low frequency of certain conditions and lab tests. Bigger samples are needed to further explore this aspect. Although the datasets share very similar qualities, they are not completely identical. This is was a positive findings as it meant that the GAN did not replicate the real seal dataset values, which would compromise privacy. Nevertheless, the degree of difference between the two datasets will need to be justified and accepted for the digital health application the RSD is intended to validate. Further validation from the point of view of clinical conditions would provide additional significant evidence on the equivalence of the two datasets. This would allow testing specific applications, and provide opportunities for expert (clinical) review of the dataset. Validation of applications using ML-generated RSDs heavily depends on contextual information about the application, as well as the generation of the dataset. A justification of use of the RSD is necessary, as it will allows the RSD developers to understand the safety implications of the generation process. Consequently, evidence need to be identified, supporting the justification. This will alleviate implications, offering evidence about fitness of the approach as a means of validation. One positive aspect about this approach is that it does not need to be developed for a specific dataset. Other approaches need statistical models (e.g., Bayesian networks) to model the real dataset, which then use it to generate the synthetic dataset. In contrast, the GAN-based RSDGM does not need to model the real dataset. This allows to have a core approach that can be applied for multiple datasets. This was confirmed during the experiments, during which the GAN successfully generated realistic synthetic datasets based on three different datasets. An additional advantage of this is that the approach maintains associations in the data that may not be yet understood, and hence modelled manually. This can be particularly helpful for datasets used for discovery of new clinical knowledge. One identified challenge of the approach is the need for significant computing power to train the network and generate the dataset. However, this is not considered prohibitive. Future work of the justification includes further developing the arguments, also identifying specific evidence for the RSD based on a proof of concept. Future work will need to focus on further validation tests of the two datasets, as well as systematically testing various configurations and hyper-parameters. Finally, different architectures of the GAN can be explored by implementing other machine learning techniques such as SVMs for the discriminator, or auto-encoders for the generator. Overall, the GAN based RSDGM showed much promise and is considered a viable approach to be used for development of a healthcare dataset.

## Data Availability

Dataset used available at MIMIC website. Data generated cannot be shared as they are part of ongoing further work in the involved organizations.

https://mimic.physionet.org/

## ACKNOWLEDGEMENTS

This work was led by the MHRA and performed in collaboration with the NHS Digital and a2-ci under the £10m Regulators’ Pioneer Fund, launched by The Department for Business, Energy and Industrial Strategy (BEIS). The fund enables UK regulators to develop innovation-enabling approaches to emerging technologies and unlock the long-term economic opportunities identified in the government’s modern Industrial Strategy. HDR UK is funded by the UK Medical Research Council, Engineering and Physical Sciences Research Council, Economic and Social Research Council, Department of Health and Social Care (England), Chief Scientist Office of the Scottish Government Health and Social Care Directorates, Health and Social Care Research and Development Division (Welsh Government), Public Health Agency (Northern Ireland), British Heart Foundation and Wellcome Trust. This is a research product and does not reflect future policies nor recommendations of the involved organisations. Special thanks to Dr Eda Bilici Ozyigit for her technical work, as a research associate on the project.

